# Prevalence and correlates of intimate partner violence among PrEP-eligible men and women in Coastal Kenya

**DOI:** 10.64898/2026.03.30.26349739

**Authors:** Tanmay Bhanushali, Liying Wang, Fred Ogadah, Elizabeth Wahome, Clara Agutu, Elise M. van der Elst, Eduard J. Sanders, Susan M. Graham

**Author notes:** Corresponding author (TB).

## Abstract

**Background:** Pre-exposure prophylaxis (PrEP) is an effective HIV prevention tool, yet uptake and adherence remain low in Kenya despite integration into national HIV prevention plans since 2017. Intimate partner violence (IPV) is a prevalent HIV-related syndemic that presents barriers to PrEP engagement. While IPV’s impact on women’s PrEP use has been documented, less is known about IPV prevalence among men and its association with PrEP eligibility. This study aimed to determine IPV prevalence and explore correlates among PrEP-eligible men and women in coastal Kenya.

**Methods:** This secondary analysis used data from the "Tambua Mapema Plus" trial conducted at six healthcare facilities in coastal Kenya among HIV-negative participants who were sexually active in the last 6 weeks and PrEP-eligible based on Kenya’s Rapid Assessment Screening Tool. IPV was assessed through screening questions covering physical, verbal, and sexual violence experiences. Participants with ongoing IPV were excluded for safety. Among 1,500 intervention participants, 638 (402 women, 236 men) met PrEP eligibility criteria. Modified Poisson regression with robust standard errors was used to identify factors associated with IPV.

**Results:** Overall, 24.1% reported lifetime IPV exposure, with 5.6% reporting past-month IPV. Women experienced higher rates of verbal (14.9% vs 11.0%), physical (15.2% vs 9.7%), and sexual IPV (11.2% vs 6.4%). Participants who had children (adjusted risk ratio [ARR]=2.09, 95%CI 1.32‒3.32) or engaged in sex work (ARR=1.81, 95%CI 1.13‒2.80) had increased IPV risk. In multivariable analysis, women with children had higher IPV risk (ARR=2.30, 95%CI 1.29‒4.24), while men engaging in sex work had elevated risk (ARR=2.37, 95%CI 1.15‒4.68).

**Discussion:** IPV prevalence was substantial. Sex work emerged as a risk factor for both sexes, while having children increased risk among women. High IPV prevalence among PrEP-eligible individuals underscores the need for integrated IPV risk assessment in PrEP programs to improve HIV prevention effectiveness in Kenya.

## INTRODUCTION

HIV remains a public health challenge worldwide, with an estimated 1.3 million new HIV infections in 2023.^1^ HIV incidence is especially high in Kenya, where according to the National Syndemic Diseases Control Council, there were 15,642 new HIV infections among adults aged 15 and over in 2025, of whom 11,033 (70.5%) were women and 4,609 (29.5%) were men. ^2^ Ongoing efforts are needed to end the HIV epidemic in Kenya.

Pre-exposure prophylaxis (PrEP) is an effective tool for HIV prevention. In 2017, Kenya integrated PrEP into its national HIV prevention plan; unfortunately, PrEP uptake and adherence have been suboptimal. One study of PrEP-eligible women found that of the 6.6% of participants who had used PrEP in the first year of the study, fewer than half persisted in PrEP use during follow-up visits.^3^ In a PrEP monitoring study involving women in Kenya aged 18-24 at high risk for HIV, a high average PrEP adherence (5+ doses per week) was observed at only 20% of participant visits.^4^ In an analysis of data from the SEARCH (Sustainable East Africa Research in Community Health) study of community residents in Kenya and Uganda, only 27% of residents with high HIV risk initiated PrEP.^5^ It is clear that although PrEP is a highly effective HIV prevention tool, low uptake and adherence limit its efficacy.

Intimate partner violence (IPV) is a highly prevalent HIV-related syndemic in Kenya and presents a significant barrier to seeking and engaging in HIV prevention services.^6^ According to a cross-sectional study of women with HIV living in Nairobi, Kenya, 15%-20% reported experiencing some form of IPV in the six months before completing the IPV questionnaire. IPV is associated with elevated risk for HIV among female survivors in the short term due to forced sex and in the longer term due to mental health sequelae and reduced safe sex behaviors.^7,8^ In the context of IPV, concerns about a partner’s reaction to PrEP use can serve as a deterrent for individuals seeking to initiate PrEP. For example, a study conducted among pregnant Kenyan women at risk for HIV found that these women believed their male partners would become physically violent if they were found to be using PrEP.^9^ Less is known about the prevalence of IPV among men and its association with PrEP eligibility and use. A review on male IPV experiences found that there is a lack of data on IPV help-seeking processes among male survivors, along with a lack of training for frontline healthcare professionals on helping male IPV victims.^10^

This is a secondary analysis using data from our completed study, the “Tambua Mapema Plus” (TMP) trial, which was conducted in coastal Kenya between 2017 and 2020.^11,12^ We assessed the impact of a targeted HIV-1 testing intervention among adults aged 18–39 years at six healthcare facilities using a modified stepped-wedge trial design. The intervention involved opt-out point-of-care HIV-1 nucleic acid amplification testing, followed by rapid antibody tests to differentiate acute from chronic infections, compared to standard provider-initiated rapid testing.^11^ We found that routine opt-out testing diagnosed twice as many individuals as discretionary testing, yielding one chronic HIV case per 40 patients and one acute HIV infection (AHI) per 750 patients tested.^12^ The current analysis aimed to determine the prevalence of IPV and explore its correlates among PrEP-eligible men and women who participated in the TMP HIV testing intervention trial in coastal Kenya.

## METHODS

### Study setting

The TMP trial was conducted in six facilities in coastal Kenya between 2017 and 2020.^11^ Each facility had an observation and intervention phase, with each phase lasting 6 months, except for the first facility, which only had 3 months of observation.^11^ In total, 2,874 participants enrolled in the study, among whom 1,374 participated in the observation phase and 1,500 participated in the intervention phase.^11^

The TMP study had the following inclusion criteria: 18-39 years of age; no previous HIV diagnosis; a score of 2 or greater on a published AHI risk score algorithm.^13^ The algorithm has the following scoring criteria: aged 18–29 years (1 point), fever (1 point), fatigue (1 point), body pains (1 point), diarrhea (1 point), sore throat (1 point), and genital ulcer disease (GUD) (3 points).^11^ Individuals who met these criteria were screened for IPV prior to enrolling in the study as described below. The TMP study was approved by the Ethical Review Committee at the Kenya Medical Research Institute (SERU protocol 3280) and the Human Subjects Division at the University of Washington (Study00001808). Participants were recruited between 1 December 2017 through 17 March 2020. All participants provided written informed consent.

### IPV screening

IPV was assessed during eligibility screening by asking participants to respond to three questions: (1) Have you ever been in a relationship with anyone who physically hurt you?; (2) Have you ever been in a relationship with a person who threatened, frightened, insulted, or treated you badly?; and (3) Have you ever been in a relationship with a person who forced you to participate in sexual activities that made you feel uncomfortable? Individuals who answered “Yes” to any of these questions were asked if these incidents were ongoing, had occurred in the past month or had occurred more than one month ago. Individuals experiencing ongoing IPV were excluded from the trial to protect participants’ safety and were referred for counseling.^11^ Individuals with more remote (i.e., past month or longer ago) experiences of IPV and those who reported no IPV were offered enrollment in the HIV testing intervention phase. After enrollment, participants with a new HIV diagnosis during the study were followed up for linkage to care and provided ongoing counseling and monitoring for IPV events.

### PrEP eligibility

This secondary data analysis used data from the subset of participants who were PrEP eligible. PrEP eligibility criteria included: (1) HIV-negative status; (2) sexually active in the 6 weeks prior to study enrollment; and (3) meeting at least one HIV risk criteria. The HIV risk criteria were based on the Kenya National AIDS and STI Control Program Rapid Assessment Screening Tool (RAST),^14^ which asks whether individuals have had any of the following in the past 6 months: (1) one or more sexual partners living with HIV, (2) sex without a condom with a partner of unknown or positive HIV status, (3) sex in exchange for money or other favors, (4) a genital ulcer disease diagnosis, (5) needle sharing while engaging in injection drug use. Use of HIV post-exposure prophylaxis in the past 6 months is a sixth criterion in the RAST, but this information was not collected in the TMP study.

TMP surveys included questions on partners living with HIV, sex without a condom with a partner of unknown or positive HIV status, sex in exchange for money or favors, and needle sharing in the past 6 weeks. In addition, data on having same-sex partners in the past 6 weeks were collected in TMP and so were included as an additional HIV risk factor for the purpose of this analysis. The only STI data collected in TMP was the presence of a genital ulcer at screening; therefore, this variable was used as a proxy for having been diagnosed with or treated for an STI.

### Demographic characteristics

Data collected on demographic characteristics in the TMP trial included age, sex, marital status, education level, religion, employment status, and whether or not participants had children.

### Data analysis

Descriptive statistics were used to compare participant characteristics and IPV prevalence in the study population, overall and stratified by sex. Chi-square tests were used to calculate p-values for differences by sex.

Modified Poisson regression with robust standard errors was used to model factors associated with IPV among PrEP-eligible TMP participants and obtain risk ratios (RR) and adjusted risk ratios (ARR) with 95% confidence intervals (CI). We conducted analysis in the overall PrEP-eligible population first, then repeated these analyses stratified by sex. As these analyses were exploratory, we did not adjust the p-value for multiple comparisons.^15^ Analysis was conducted using R version 4.2.2.

## RESULTS

### Participant characteristics

Of the original 1500 participants in the intervention group, 638 participants (402 women and 236 men) met the PrEP eligibility criteria detailed above and were included in this study. Participants were young, with 271 (42.5%) aged 18-24 (Table 1). Most were married (56.6%) and Christian (78.1%), while almost half (46.7%) had not completed secondary education. In terms of risk factors, 37 participants (5.8%) reported engaging in transactional sex in the past 6 weeks, while only 4 (0.6%) reported same-sex partners and 1 (0.2%) reported sharing needles while engaged in injection drug use. About 13.8% (n=88) of participants reported a genital ulcer at the TMP study visit. Table 1 also compares characteristics of male and female participants. Women were less likely to be employed than men (46.0% versus 67.4%) and were less likely to have attained higher education than men (15.7% versus 30.5%). Women were more likely than men to have children (73.1% versus 46.2%, respectively) and to be married (64.9% versus 42.4%, respectively).

**Table 1.**
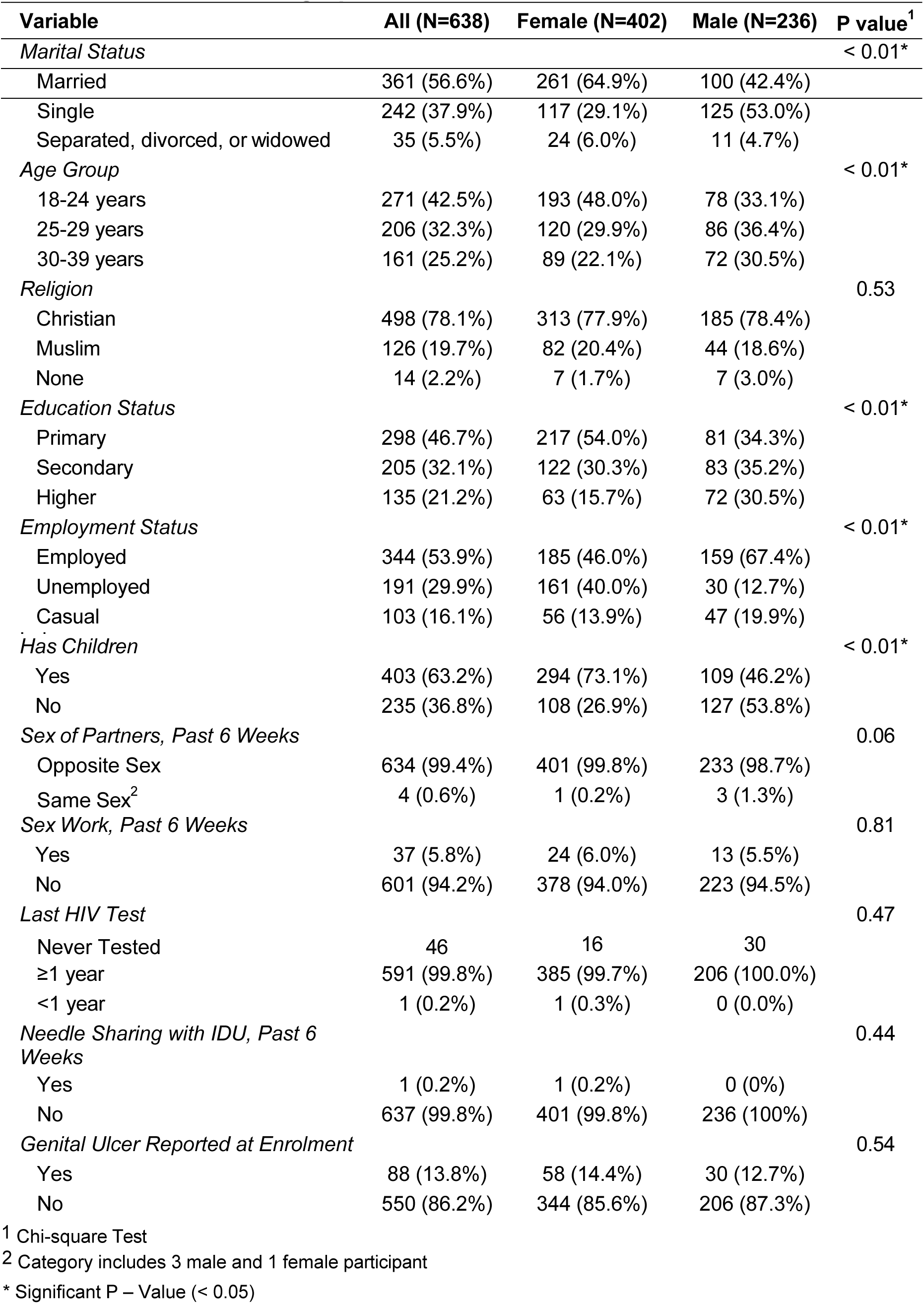
Social and Demographic Characteristics.

Table 2 presents the prevalence of different forms of IPV and IPV overall. Approximately 24.1% (n=154) of participants had experienced some type of IPV over their lifetime, with 5.6% (n=34) of participants reporting IPV in the last month. Female participants had experienced each type of IPV at higher rates than male participants: 14.9% versus 11.0% for verbal IPV, 15.2% versus 9.7% for physical IPV, and 11.2% versus 6.4% for sexual IPV. This difference was the only statistically significant for sexual IPV experiences (*p* = 0.04). The most common form of IPV among female participants was physical IPV (15.2%), while male participants most commonly reported verbal IPV (11.0%).

**Table 2.** IPV Prevalence.

| Variable | All (N=638) | Female (N=402) | Male (N=236) | P Value<sup>1</sup> |
| --- | --- | --- | --- | --- |
| Any Physical IPV | 84 (13.2%) | 61 (15.2%) | 23 (9.7%) | 0.05* |
| Any Verbal IPV | 86 (13.5%) | 60 (14.9%) | 26 (11.0%) | 0.16 |
| Any Sexual IPV | 60 (9.4%) | 45 (11.2%) | 15 (6.4%) | 0.04* |
| Any IPV | 154 (24.1%) | 107 (26.6%) | 47 (19.9%) | 0.06 |
| Recent<sup>2</sup> Physical IPV | 16 (2.5%) | 7 (1.7%) | 9 (3.8%) | 0.11 |
| Recent<sup>2</sup> Verbal IPV | 17 (2.7%) | 10 (2.5%) | 7 (3.0%) | 0.72 |
| Recent<sup>2</sup> Sexual IPV | 12 (1.9%) | 10 (2.5%) | 2 (0.8%) | 0.14 |
| Any Recent<sup>2</sup> IPV | 34 (5.6%) | 19 (5.0%) | 15 (6.6%) | 0.41 |

<sup>1</sup> Chi-square Test
<sup>2</sup> "Recent" was defined as occurring in the past month.
\* Significant P – Value (< 0.05)

### Correlates of IPV among all participants

Table 3 presents results of the analysis of correlates of IPV in the entire study sample. In bivariable analysis, participants who were separated, divorced, or widowed had a higher risk of having experienced IPV (RR = 1.96, 95%CI 1.11‒3.28) compared to participants who were single. Participants with children also had a higher risk of having experienced IPV in comparison to participants without children (RR = 1.51, 95%CI 1.07‒2.16). Participants who reported engaging in sex work had a higher risk of having experienced IPV compared to those who did not (RR = 2.17, 95%CI 1.40‒3.23). In multivariable analysis, participants who had children (ARR=2.09, 95%CI 1.32‒3.32) or engaged in sex work (ARR=1.81, 95%CI 1.13‒2.80) had increased IPV risk.

**Table 3.** Factors associated with any IPV among all participants eligible for PrEP (N=638)

| Variable | Bivariable |  |  | Multivariable |  |  |
| --- | --- | --- | --- | --- | --- | --- |
|  | RR | 95% CI | P value<sup>1</sup> | RR | 95% CI | P value<sup>1</sup> |
| Marital Status |  |  | 0.02* |  |  | 0.06 |
| Single | Reference |  |  | Reference |  |  |
| Separated, divorced, or widowed | 1.96 | 1.11 – 3.28 |  | 1.37 | 0.71 – 2.56 |  |
| Married | 0.86 | 0.61 – 1.21 |  | 0.72 | 0.47 – 1.13 |  |
| Has Children | 1.51 | 1.07 – 2.16 | 0.02* | 2.09 | 1.32 – 3.32 | 0.01* |
| Education Status |  |  |  |  |  |  |
| Primary | Reference |  | 0.74 | Reference |  | 0.36 |
| Secondary | 1.14 | 0.79 – 1.62 |  | 1.24 | 0.84 – 1.83 |  |
| Higher | 0.99 | 0.64 – 1.50 |  | 1.24 | 0.77 – 1.97 |  |
| Same-sex partners<sup>2</sup> | NC** | NC | NC | 1.81 | 0.29 – 6.06 | 0.42 |
| Sex Work | 2.17 | 1.40 – 3.23 | 0.01* | 1.81 | 1.13 – 2.80 | 0.01* |
| Age Group |  |  |  |  |  |  |
| 18-24 Years Old | Reference |  | 0.81 | Reference |  | 0.38 |
| 25-29 Years Old | 0.89 | 0.61 – 1.29 |  | 0.78 | 0.52 – 1.16 |  |
| 30–39 Years Old | 0.99 | 0.66 – 1.46 |  | 0.77 | 0.49 – 1.19 |  |
| Employment Status |  |  |  |  |  |  |
| Unemployed | Reference |  | 0.87 | Reference |  | 0.36 |
| Employed | 1.05 | 0.73 – 1.52 |  | 1.05 | 0.72 – 1.55 |  |
| Casual Laborer | 1.14 | 0.70 – 1.82 |  | 1.12 | 0.67 – 1.84 |  |

<sup>1</sup> Chi-square Test
<sup>2</sup> Category includes 3 male and 1 female participant
\* Significant P Value (< 0.05)
\*\*NC = No Convergence

### Correlates of IPV stratified by gender

Table 4 presents the analysis restricted to female participants. Among female participants, the bivariable Poisson regression showed that women who reported engaging in sex work (RR = 2.28, 95%CI 1.25‒3.87), women with children (RR = 1.71, 95%CI 1.07‒2.89), and women who were separated, divorced, or widowed (RR = 2.02, 95%CI 1.05‒3.69]) had a higher risk of having experienced IPV, in comparison to those who did not engage in sex work, did not have children, and were not separated, divorced, or widowed. In multivariable analysis, women with children had a higher risk of having experienced IPV compared to those without children (ARR = 2.30, 95%CI 1.29‒4.24), after controlling for other variables in the model.

**Table 4.** Factors associated with any IPV among female participants eligible for PrEP (N=402)

| Variable | Bivariable |  |  | Multivariable |  |  |
| --- | --- | --- | --- | --- | --- | --- |
|  | RR | 95% CI | P value<sup>1</sup> | RR | 95% CI | P value<sup>1</sup> |
| Marital Status |  |  | 0.01* |  |  | 0.06 |
| Single | Reference |  |  | Reference |  |  |
| Separated, divorced, or widowed | 2.02 | 1.05 – 3.69 |  | 1.55 | 0.74 – 3.14 |  |
| Married | 0.78 | 0.52 – 1.20 |  | 0.71 | 0.43 – 1.19 |  |
| Has Children | 1.71 | 1.07 – 2.89 | 0.02* | 2.30 | 1.29 – 4.24 | 0.01* |
| Education Status |  |  |  |  |  |  |
| Primary | Reference |  | 0.38 | Reference |  | 0.16 |
| Secondary | 1.29 | 0.84 – 1.95 |  | 1.52 | 0.96 – 2.39 |  |
| Higher | 1.11 | 0.62 – 1.88 |  | 1.50 | 0.80 – 2.66 |  |
| Same-sex partners<sup>2</sup> | NC** | NC | NC | NC | NC | NC |
| Sex Work | 2.28 | 1.25 – 3.87 | 0.01* | 1.70 | 0.89 – 3.06 | 0.11 |
| Age Group |  |  |  |  |  |  |
| 18-24 Years Old | Reference |  | 0.52 | Reference |  | 0.38 |
| 25-29 Years Old | 0.88 | 0.54 – 1.35 |  | 0.71 | 0.43 – 1.15 |  |
| 30–39 Years Old | 1.11 | 0.69 – 1.76 |  | 0.88 | 0.53 – 1.44 |  |
| Employment Status |  |  |  |  |  |  |
| Unemployed | Reference |  | 0.42 | Reference |  | 0.65 |
| Employed | 1.26 | 0.83 – 1.93 |  | 1.21 | 0.78 – 1.88 |  |
| Casual Laborer | 1.22 | 0.66 – 2.16 |  | 1.24 | 0.66 – 2.24 |  |

<sup>1</sup> Chi-square Test
<sup>2</sup> Category includes 3 male and 1 female participant
\* Significant P Value (< 0.05)
\*\*NC = No Convergence

Table 5 presents the analysis restricted to male participants. Among male participants, bivariable analysis showed that men who reported engaging in sex work (RR = 2.41, 95%CI 1.23‒4.46) had a higher risk of having experienced IPV compared to those who did not engage in sex work. In the multivariable analysis, this association remained significant (RR = 2.37, 95%CI 1.15‒4.68), after controlling for other variables in the model.

**Table 5.** Factors associated with any IPV among male participants eligible for PrEP (N=236)

| Variable | Bivariable |  |  | Multivariable |  |  |
| --- | --- | --- | --- | --- | --- | --- |
|  | RR | 95% CI | P value<sup>1</sup> | RR | 95% CI | P value<sup>1</sup> |
| Marital Status |  |  | 0.78 |  |  | 0.95 |
| Single | Reference |  |  | Reference |  |  |
| Separated, divorced, or widowed | 1.30 | 0.31 – 3.69 |  | 1.08 | 0.21 – 4.25 |  |
| Married | 0.87 | 0.47 – 1.57 |  | 0.90 | 0.35 – 2.38 |  |
| Has Children | 1.12 | 0.63 – 1.99 | 0.67 | 1.31 | 0.53 – 3.13 | 0.56 |
| Education Status |  |  |  |  |  |  |
| Primary | Reference |  | 0.97 | Reference |  | 0.90 |
| Secondary | 0.91 | 0.42 – 1.91 |  | 0.89 | 0.43 – 1.84 |  |
| Higher | 0.93 | 0.41 – 2.04 |  | 1.05 | 0.47 – 2.31 |  |
| Same-sex partners<sup>2</sup> | 2.56 | 0.42 – 8.26 | 0.26 | 2.27 | 0.35 – 8.39 | 0.34 |
| Sex Work | 2.41 | 1.23 – 4.46 | 0.01* | 2.37 | 1.15 – 4.68 | 0.02* |
| Age Group |  |  |  |  |  |  |
| 18-24 Years Old | Reference |  | 0.90 | Reference |  | 0.75 |
| 25-29 Years Old | 1.03 | 0.53 – 2.05 |  | 1.05 | 0.48 – 2.26 |  |
| 30–39 Years Old | 0.89 | 0.42 – 1.85 |  | 0.77 | 0.29 – 2.01 |  |
| Employment Status |  |  |  |  |  |  |
| Unemployed | Reference |  | 0.64 | Reference |  | 0.58 |
| Employed | 0.93 | 0.47 – 1.85 |  | 0.68 | 0.28 – 1.83 |  |
| Casual Laborer | 0.94 | 0.46 – 1.90 |  | 0.91 | 0.32 – 2.74 |  |

<sup>1</sup> Chi-square Test
<sup>2</sup> Category includes 3 male and 1 female participant
\* Significant P Value (< 0.05)

## DISCUSSION

This study examined the prevalence and correlates of IPV among PrEP-eligible men and women in coastal Kenya, a population situated at the intersection of HIV vulnerability and gendered power relations. We found that 24.1% of PrEP-eligible participants reported lifetime IPV exposure, with women experiencing higher rates of each type of violence than men.

Engaging in sex work and having children were associated with increased risk for IPV among all participants, with having children emerging as the strongest predictor of IPV for female participants and sex work as the strongest predictor of IPV for male participants.

Lifetime exposure to any form of IPV was 19.9% and 26.6% among male and female participants, respectively. This is lower than the female IPV rates found in a study of Kenya National Demographic and Health Survey (KDHS) data by Melkam and colleagues, which reported a lifetime IPV prevalence of 41.1% for female respondents aged 15-49, with 29.9% experiencing physical IPV, 27.8% experiencing verbal IPV, 10.7% experiencing economic violence, and 10.3% experiencing sexual IPV.^16^ In our analysis, female TMP participants reported less physical and verbal violence (15.2% and 14.9%, respectively), but more sexual IPV, at 11.2%. These differences could be partially due to narrower age range used in the TMP study, which focused on women aged 18-39, and the exclusion of economic violence from the IPV definition we used. In addition, our study excluded women who reported ongoing IPV at screening, biasing our results towards a lower estimate. Importantly, while ethically appropriate for participant safety, this exclusion may have resulted in a survivor-selection effect, whereby individuals most acutely impacted by IPV – and potentially facing the greatest barriers to PrEP uptake and adherence – were underrepresented in the analytical sample.

IPV prevalence among males in our study population was noticeably lower than that among females. This is consistent with a study by Mukthar et al that also used KDHS data and found that 19.6% of males experienced verbal IPV, 3.1% experienced physical IPV, and 3.7% experienced sexual IPV.^17^ Of interest, research among male-male couples has found that men tend to under-report IPV experiences,^18^ and societal stigma against male-male sex could have led to an underestimation of IPV rates among such men in our study. This stigma can also extend to male experiences of participating in commercial sex work. Among heterosexual men in Kenya, underreporting may also occur, due to factors such as stigma associated with seeking help and societal concepts of traditional masculinity.^19^ More research on IPV among males in Kenya, especially among men eligible for PrEP, is needed to better understand men’s experiences, how they influence reporting and help-seeking, and implications for HIV prevention.

Factors influencing IPV prevalence differed between the genders as well. We found that female participants with children were more likely to experience IPV. Two other studies conducted in Kenya have identified multiparity as a predictor of IPV prevalence among women.^20,21^ In this context, having children may reflect broader social and economic dynamics, including caregiving responsibilities, economic dependency, and reduced capacity to leave unsafe relationships. Sociocultural norms including a high tolerance for domestic violence against women and marital customs such as the payment of dowry could also factor in.^22^ Research to develop culturally appropriate interventions to protect women with children from IPV remains imperative.

Sex work was a risk factor for increased IPV prevalence for both female and male participants in bivariable analysis, with a similar estimated relative risk (2.3 and 2.4, respectively). IPV is highly prevalent among sex workers in Kenya, with prior studies documenting high levels of physical and sexual violence.^23^ Factors such as high rates of alcohol use at sex work venues, partner or client control, stigma, and criminalisatation are thought to contribute to this elevated risk.^24^ Because unhealthy alcohol use among Kenyan women who engage in sex work has been associated with sexual IPV, the integration of IPV and alcohol treatment services has been recommended for this population.^25^ The association observed among men highlights a potentially hidden vulnerability, as male sex workers may face compounded stigma and barriers assessing support services.^26^ Of note, engaging in sex work may decrease PrEP acceptability and uptake among both men and women. For example, transactional sex was correlated with decreased PrEP adherence in a study of men who have sex with men in coastal Kenya.^27^ Additionally, over longitudinal follow-up of women engaged in sex work in the Kyaterekera study in Uganda, investigators found that a longer duration of sex work was associated with lower willingness to use PrEP.^28^ Screening for sex work engagement may therefore assist providers in identifying individuals at risk for IPV and providing appropriate support and HIV risk reduction counseling.

Although not examined in this study, prior research has demonstrated that verbal, physical, and economic IPV are correlated with low PrEP adherence in Kenyan women.^29^ Qualitative data suggest mechanisms including IPV-related stress, partner interference, and difficulties with medication storage or disclosure.^29^ These findings support conceptualizing IPV as a factor influencing engagement across the PrEP care continuu, rather than solely a correlate of HIV risk.

The current HIV prevention landscape also warrants consideration. Oral PrEP was the only modality available during the TMP study period. Emerging long-acting PrEP formulations may offer advantages for individuals experiencing IPV by increasing discretion, and reducing the need for daily pill-taking. Research is needed to understand the extent to which IPV impacts the PrEP care continuum for women and men who experience IPV, as well as for individuals engaging in sex work or in same-sex partnerships. Given the negative impact of IPV on PrEP uptake and adherence, targeted IPV risk assessment before a prescription for oral PrEP (the only type of PrEP currently available in Kenya) is warranted. However, bio-medical advances alone are insufficient. IPV screening within PrEP programs should be accompanied by referral pathways, psychological support, and provider training to ensure that identification of IPV does not inadvertently increase risk.

This study has several limitations. First, the TMP study was conducted in a limited geographical area, restricting generalizability. Second, IPV and sexual behavior were self-reported. Third, IPV screening excluded individuals experiencing ongoing IPV, limiting participation to those with no or limited IPV experiences. As noted above, this exclusion likely resulted in conservative prevalece estimates and under-representation of those most vulnerable to poor PrEP outcomes. Fourth, the questions and recall period for data collected for the TMP study did not align perfectly with Kenya’s RAST tool; since the RAST tool’s recall period is longer, this may have led to some TMP participants who would be eligible for PrEP being misclassified as ineligible. Fifth, limited numbers of participants reporting same-sex relationships precluded sub-group analyses. Nevertheless, this study included a large population of both men and women in the same communities in coastal Kenya, enabling us to compare these groups and provide significant results. We have been able to estimate the prevalence of different types of IPV among Kenyan men and women in the study area and describe how HIV risk behaviors, such as transactional sex, correlate with past experiences of IPV.

In conclusion, this study is one of only a few studies that have analyzed IPV prevalence and its correlates among PrEP-eligible individuals in Kenya. We found substantial IPV prevalence among women and lower but still concerning IPV prevalence patterns among men, supporting the need for more research studying IPV experiences in both sexes. Sex work and having children were associated with IPV experiences, underscoring the importance of considering structural and relational vulnerabilities within HIV prevention programs. Integrating IPV assessment with supportive, trauma-informed PrEP services may be essential to improving HIV effectiveness in Kenya.

## Data Availability

Deidentified data and code for this analysis will be deposited in the Dryad Data Repository upon acceptance of the final version.

## Acknowledgements

We thank staff at the six health facilities in Kilifi and Mombasa counties for their support, study participants for their commitment to the study, and staff at the KEMRI-Wellcome Trust Programme for facilitating the conduct of the TMP trial.

## Notes

### Competing Interest Statement

The authors have declared no competing interest.

### Clinical Trial

The TMP trial is registered with ClinicalTrials.gov NCT03508908.

### Author Declarations

The TMP trial received ethical approval by the KEMRI Scientific and Ethical Review Unit (KEMRI/SERU/CGMRC-C/051/3280), the Human Subjects Division at the University of Washington (STUDY00001808), and the Oxford Tropical Research Ethics Committee (OxTREC) at the University of Oxford (reference: 46-16). All participants provided written informed consent.

